# Using ^18^F-NOS PET Imaging to Measure Pulmonary Inflammation in Electronic and Combustible Cigarette Users: A Pilot Study

**DOI:** 10.1101/2022.06.15.22276438

**Authors:** Reagan R. Wetherill, Robert K. Doot, Anthony J. Young, Hsiaoju Lee, Erin K. Schubert, Corinde E. Wiers, Frank T. Leone, Robert H. Mach, Henry R. Kranzler, Jacob G. Dubroff

**Author notes:** Corresponding Author: Reagan R. Wetherill, Ph.D., 3535 Market Street, Suite 500, Philadelphia, PA 19104.

## Abstract

Electronic cigarette (EC) use has increased dramatically, particularly among adolescents and young adults, which, like cigarette use, can cause inflammation of the lungs and increase the risk of lung disease.

**Methods:** In this preliminary study, we used positron emission tomography with ^18^F-6-(1/2)(2-fluoro-propyl)-4-methylpyridin-2-amine (^18^F-NOS) to quantify inflammation of the lungs *in vivo* in three age- and sex-matched groups: (1) 5 daily EC users, (2) 5 daily cigarette smokers, and (3) 5 never smoke/vape controls.

**Results:** EC users showed greater ^18^F-NOS non-displaceable binding potential (BP_ND_) than cigarette smokers (*p* = 0.03) and never smoke/vape controls (*p* = 0.01); whereas BP_ND_ in cigarette smokers did not differ from controls (p > 0.1). ^18^F-NOS lung tissue delivery (K_1_) and iNOS distribution volume (V_T_) did not significantly differ between groups. Although there were no group differences in the concentration of the peripheral inflammatory markers TNF-α, IL-6 or IL-8, ^18^F-NOS BP_ND_ significantly correlated with the proinflammatory cytokine TNF-α (*r* = 0.87, *p* = 0.05) in EC users. Additionally, when EC users and cigarette smokers were pooled together, vaping episodes/cigarettes per day correlated with IL-6 levels (*r* = 0.86, *p* = 0.006).

**Conclusion:** To our knowledge, this is the first PET imaging study to compare lung inflammation between EC and cigarette users *in vivo*. We found preliminary evidence EC users had greater pulmonary inflammation than cigarette smokers and never smoke/vape controls, with a positive association between pulmonary and peripheral measures of inflammation.

## Introduction

Tobacco use is the world’s greatest preventable cause of morbidity and mortality, accounting for more than 8 million deaths each year (*1*). Although public awareness of smoking-related risks has increased and tobacco smoking has declined in many countries, electronic cigarette (EC) use has increased dramatically, particularly among youth and young adults (*1*–*3*). This increase in EC use is partially driven by the assumption that ECs are safer than conventional cigarettes. Although ECs are often advertised as an alternative smoking cessation tool (*4,5*), their long-term effectiveness and safety have not been rigorously evaluated (*6,7*). Given the emergence of the EC or Vaping Product-Associated Lung Injury (EVALI) epidemic in 2019 (*8*), EC use has become a major public health concern and the adverse pulmonary effects of EC use, particularly relative to that of cigarette smoking, remain unclear.

ECs deliver nicotine by heating e-liquids containing nicotine in a vegetable glycerin/propylene glycol vehicle with flavorings that are vaporized and inhaled, thus delivering nicotine without combusting tobacco. Although the propylene glycol and vegetable glycerin found in e-liquids are commonly used in food and cosmetic products and are regarded as “safe” by the Food and Drug Administration (FDA), e-aerosols contain tobacco-specific nitrosamines (TSNAs), metals, polycyclic aromatic hydrocarbons (PAHs), and volatile organic compounds (VOCs) that are known toxicants and carcinogens (*9*). As with smoking, several of these EC-related compounds are associated with inflammation, an altered innate immune response, oxidative stress, and cytotoxicity (*9*–*11*). However, the existing human literature on the pulmonary effects of EC use is limited and comprised mainly of studies that use invasive approaches (e.g., induced sputum, bronchoalveolar lavage and brushings) that do not adequately assess the impact of EC use on the lungs.

Positron emission tomography (PET) imaging has been used to quantify and track inflammatory responses associated with smoking and EC use *in vivo* without the need for invasive diagnostic studies (*12,13*). PET with ^18^F-fluorodeoxyglucose (^18^F-FDG) has been used extensively to detect enhanced glucose metabolic activity of activated immune cells in inflammatory diseases, including pneumonia (*14*), cystic fibrosis (*14*), COPD (*15*), and sarcoidosis (*16*). Although associations between ^18^F-FDG quantification and inflammation have been observed, biologic processes, including fibrosis and neoplasia, utilize glucose and consequently limit the specificity of ^18^F-FDG (*17*). PET radiotracers targeting the 18 kDa translocator protein (TSPO, also known as the peripheral benzodiazepine receptor) have also been used to measure pulmonary inflammation (*18,19*). While these radiotracers were initially considered putative markers of neuroinflammation, their specificity for inflammation is limited (*20*). Thus, recent efforts have focused on imaging specific aspects of immune regulation and response, such as nitric oxide synthase (NOS) enzymes, with promising results (*17,21*).

Nitric oxide (NO) plays an important role in immune regulation (*22*) and is produced by three nitric oxide synthase (NOS) enzymes: neuronal NOS (nNOS), endothelial NOS (eNOS), and inducible NOS (iNOS). iNOS is associated with acute and chronic inflammatory diseases, including asthma and chronic obstructive pulmonary disease (COPD) (*21,23,24*), and is expressed in normal lung epithelium (*25*). There is convergent evidence that iNOS plays a central role in mediating inflammation in combustible cigarette smokers, thereby contributing to smoking-related lung diseases. Preclinical models show that chronic exposure to cigarette smoke also increases iNOS expression in brain (*26*), whereas pharmacological inhibition of iNOS reverses tobacco-induced lung disease (*27*). In humans, lung tissue from smokers with lung cancer contains higher iNOS levels than nonsmokers (*28*). Additionally, preclinical research has provided a mechanistic link between iNOS expression in the lung and inflammatory lung diseases (*27,29*). These findings strongly support iNOS as a mechanistically relevant target for molecular imaging of lung inflammation and inflammatory lung diseases.

The novel PET radiotracer ^18^F-NOS (^18^F-6-(1/2)(2-fluoro-propyl)-4-methylpyridin-2-amine) permits the visualization and measurement of *in vivo* inducible nitric oxide synthase (iNOS) expression (*17,30*). ^18^F-NOS is a radiolabeled version of a reversible iNOS inhibitor with better selectivity than other NOS enzymes (*30*). ^18^F-NOS has been validated in an animal model of lipopolysaccharide-induced lung injury (*31*) and was used successfully to image iNOS expression in humans to characterize oxidative stress and inflammation in the heart and lungs *in vivo* (*17,30*). This study used ^18^F-NOS PET lung imaging to quantify group differences in iNOS expression among EC users, cigarette smokers, and never smoke/vape controls. Based on preclinical research showing that exposure to e-liquid vapor and cigarette smoke increases iNOS expression in brain tissue (*26,32*), we hypothesized that EC users and cigarette smokers would show greater pulmonary iNOS uptake than never smoke/vape controls. Secondary study aims were to assess concentrations of peripheral biomarkers of inflammation (TNF-α, IL-6, IL-8) and to examine their association with ^18^F-NOS PET lung imaging parameters.

## MATERIALS AND METHODS

### Participants

All procedures were approved by the University of Pennsylvania Institutional Review Board and conducted in compliance with the Health Insurance Portability and Accountability Act under exploratory Investigational New Drug (IND) #140,976 for ^18^F-NOS. Written informed consent was obtained from all subjects before participation in this study. Fifteen age-and sex-matched volunteers (5 exclusive EC users [mean age=27 ± 7], 5 cigarette smokers [mean age 35± 9], and 5 never smoke/vape controls [mean age=28 ± 7]), comprising 2 women and 3 men in each group, were recruited via local print media, social media, and from among participants in previous research studies (see Table 1 for participant characteristics). All subjects underwent screening procedures that included a physical examination, medical history, routine clinical laboratory tests, and toxicologic urine analysis. Briefly, exclusion criteria included a history or evidence of significant medical disorders on history of physical examination, a lifetime DSM-5 diagnosis of a psychiatric or substance use disorder (except tobacco use disorder for EC users and cigarette smokers), a positive urine drug screen (including cannabis use), use of inhaled or oral corticosteroids or anti-inflammatory medications, and a past-month history of lung trauma or active lung infection that could impact the uptake of ^18^F-NOS. All female participants were required to have a negative pregnancy test on the scanning day before receiving the radiotracer. EC users and smokers were required to have smoked or vaped nicotine daily for the past six months. Current smoking status in the EC user and smoking group was confirmed by carbon monoxide levels greater than 10 parts per million (ppm) and urine cotinine levels greater than 150 ng/mL.

**Table 1.** Participant characteristics

|  | **Electronic**<br>**Cigarette**<br>**Users** | **Combustible**<br>**Cigarette**<br>**Users** | **Never**<br>**Smoke/Vape**<br>**Controls** | **P-value** |
| --- | --- | --- | --- | --- |
| Age (y) | 27 ± 7 | 35 ± 9 | 28 ± 7 | 0.16 |
| Sex | 3 M; 2 F | 3 M; 2 F | 3 M; 2 F | 1.00 |
| Cigarettes/day | - | 8 ± 4 | - | - |
| Vaping episodes/day | 7 ± 4 | - | - | - |
| Fagerström Test for Cigarette<br>Dependence | - | 5 ± 2 | - | - |
| Penn State E-Cigarette<br>Dependence | 6 ± 4 | - | - | - |
| HADS – Depression | 1 ± 1 | 1 ± 1 | 1 ± 1 | 0.81 |
| HADS - Anxiety | 3 ± 2 | 3 ± 2 | 1 ± 1 | 0.35 |

M = Male; F = Female; - = not applicable; HADS = Hospital Anxiety and Depression Scale.
Continuous data are mean ± SD.

Before scanning, participants completed the Hospital Depression and Anxiety Scale (*33*) to assess symptoms of depression and anxiety. EC users completed measures of vaping behavior, including the Penn State Electronic Cigarette Dependence Index (*34*), and cigarette smokers completed measures of tobacco smoking behavior, including the Fagerström Test for Cigarette Dependence (*35*). A blood sample was obtained for measurement of plasma cytokine concentrations (tumor necrosis factor-alpha (TNF-α), interleukin (IL)-6, and C-reactive protein (CRP). Participants underwent ^18^F-NOS dynamic PET/CT imaging of the thorax with venous blood sampling to determine the PET radiotracer parent fraction as a function of time for the subsequent pharmacokinetic analysis of the PET data.

### Data Acquisition

The PET radiotracer ^18^F-NOS was synthesized as previously described (*30*). Subjects were scanned with a Philips Ingenuity PET/CT scanner (Philips Healthcare, Cleveland, OH, USA), which has a 5-mm full-width at half maximum PET spatial resolution and an 18-cm axial field of view (*36*). For each study, thoracic scanning field of view to best include heart and lungs was determined by a Nuclear Medicine physician (JGD). After a low-dose attenuation-correction CT scan, a 1-h PET dynamic acquisition was started at the time of an intravenous bolus injection of ^18^F-NOS (199 ± 27 MBq) with the following framing schedule: 24 × 5-s, 6 × 10-s, 3 × 20-s, 2 × 30-s, 5 × 60-s, and 10 × 5-min frames. The attenuation-correction CT images were reconstructed into PET images using a previously described list-mode, blob-based, ordered-subsets, maximum-likelihood, expectation-maximization algorithm, including flight-time and physical-data corrections (*36*).

### Metabolite Analysis

Venous blood was sampled at approximately 2, 5, 10, 15, 30, 45, and 60 min after injection to measure radiometabolites. Activity concentrations in whole blood and plasma were counted using a WIZARD^2^ 2480 gamma counter (Perkin Elmer, Waltham, MA). Acetonitrile-treated plasma supernatant was analyzed in a 1260 Infinity Series (Agilent Technologies, Santa Clara, CA) high-performance liquid chromatology system using an Agilent ZORBAX StableBond C18 column via a mobile phase of 73% 0.1 M ammonium formate buffer and 27% methanol. The resulting plasma-to-whole blood ratio as a function of time was used to convert the image-derived whole blood input function into a plasma input function. The resulting parent PET radiotracer fraction as a function of time and the plasma input function were inputted for subsequent kinetic analysis.

### Volumes of Interest

Whole-blood pool time activity curves (TACs) were measured using 1 cm^3^ peak volumes of interest (VOIs) within 2-cm diameter spherical search VOIs within the pulmonary artery, as this blood pool is sufficiently large to minimize partial volume effects and located immediately before blood enters the lungs (Figure 1). Lung uptake TACs were extracted from all lung tissue in the PET field of view (Figure 1).

**Figure 1.**
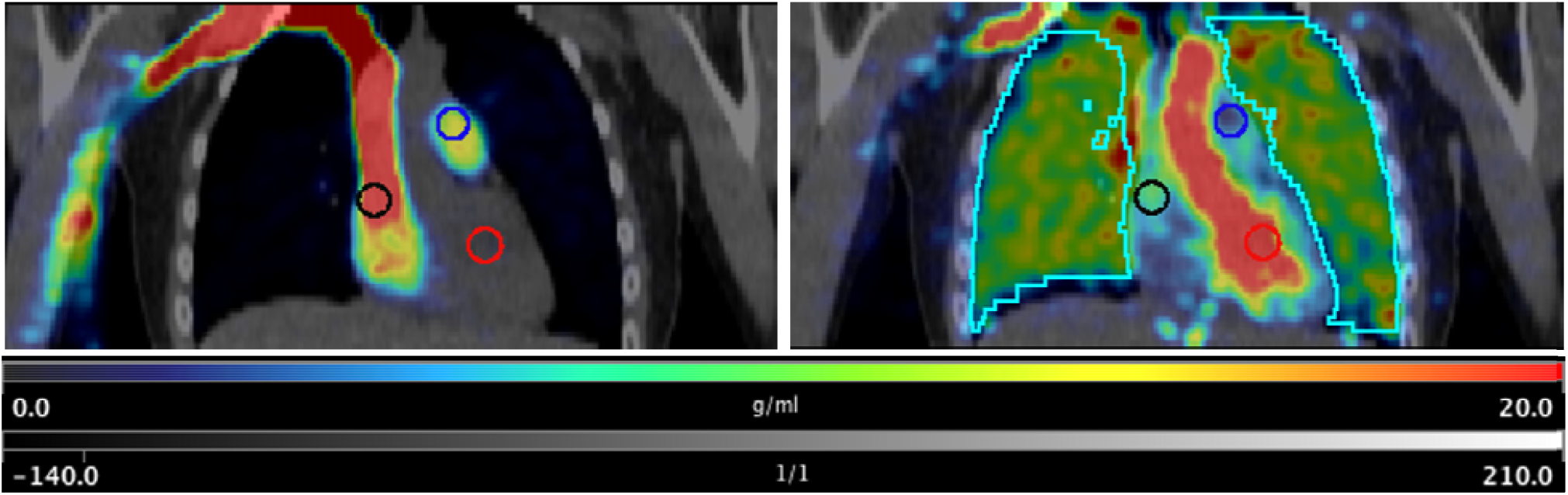
Representative fused coronal PET/CT images after injecting 207 MBq [^18^F]NOS with 2-cm diameter spherical blood pool search VOIs in right atrium (black), pulmonary artery (blue), and left ventricle (red) with PET summed uptake 0-15 s post injection in upper left panel and PET summed 37 to 42 s post injection with lung VOI (cyan) in upper right panel.

### Kinetic Analysis

Three models of kinetic analysis were compared: 1-tissue compartment (1TC), 2-tissue compartment (2TC), and Logan plot. 2TC was selected based on having the best Akaike information criteria score. Additionally, two approaches for blood volume fraction (vB) were examined for each model: fixed at 0.15 and floating between 0.05 and 0.3. Floating vB resulted in the least model variability. Kinetic analyses using a 2-compartment model with a floated lung blood volume fraction (vB) were performed to estimate total volume of distribution (V_T_), transport into first tissue compartment (K1), distribution volume of the first tissue compartment (K1/k2), and non-displaceable binding potential (BP_ND_) via Pmod image analysis software (Pmod v3.7, PMOD Technologies Ltd., Zurich, Switzerland) using the combined lung TAC and PET image-derived plasma input function from the pulmonary artery blood pool (see Figure 1). Kinetic analyses were based on the first 40 min of PET acquisition to allow a consistent analysis of all subjects’ data after one subject’s excessive motion resulted in unevaluable PET images after 40 minutes.

### Statistical Analysis

Non-parametric Mann-Whitney and Kruskal-Wallis tests were used to assess group differences. Spearman rank-order correlations measured the strength and direction of associations between biomarkers of inflammation, nicotine use behaviors (cigarettes per day and nicotine dependence for cigarette smokers; vaping episodes per day and e-cigarette dependence for EC users), and imaging parameters. All statistical tests were two-sided.

## RESULTS

Table 1 provides participant characteristics. On average, EC users reported 7 ± 4 vaping episodes/day, with Penn State Electronic Cigarette Dependence Index scores of 6 ± 4, indicating moderate-to-high levels of EC dependence. Cigarette smokers reported smoking 8 ± 4 cigarettes/day, with Fagerström Test for Cigarette Dependence scores of 5 ± 2. These characteristics are consistent with a moderate level of cigarette dependence. There were no significant group differences in age, depression or anxiety scores. There were no significant group differences on injected mass radioactivity dose or plasma-free fraction.

Selection of the pulmonary artery to measure the blood input function is supported by the example fused PET/CT images in Figure 1, where the distribution of ^18^F-NOS before entry into the lungs indicates ^18^F-NOS entering the right atrium followed by the pulmonary artery before lung entry and subsequent transport to lungs followed by the left ventricle. Average lung ^18^F-NOS uptake for all subjects as a function of time is in Figure 2. Kinetic analysis results are in Table 2, where the average estimate of lung blood volume fraction (vB) of 0.15 ± 0.02 (± SD) is consistent with the reported normal lung vB range of 0.14 to 0.19 from ^18^F-FDG PET/CT scans(*37*).

**Figure 2.**
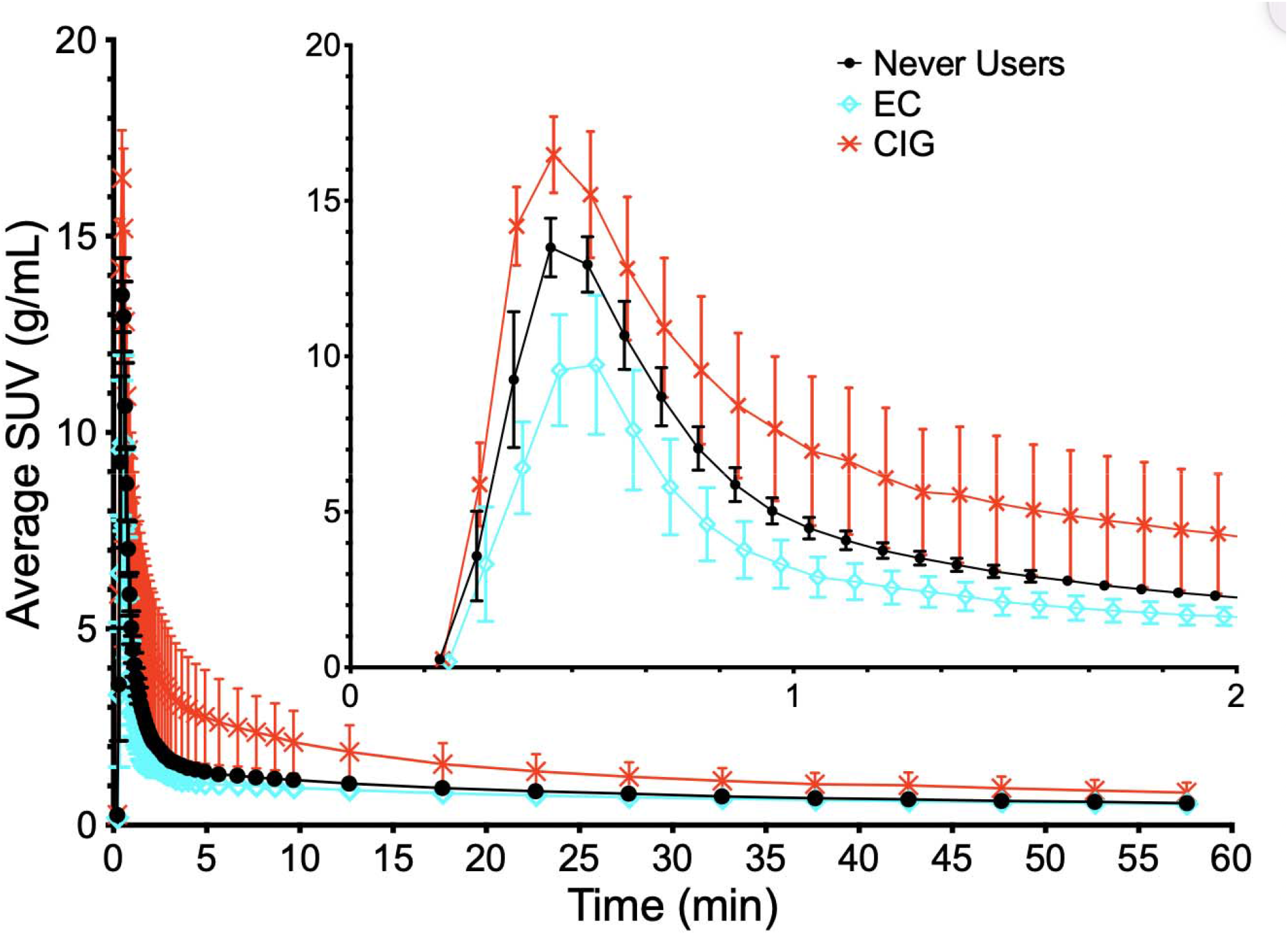
Average lung ^18^F-NOS uptake for each group as functions of time with standard deviation error bars. EC = E-Cigarette Users, CIG = Cigarette Smokers, Never Users = Never Smoke/Vape Controls.

**Table 2.** Individual and cohort gender, age, and ^18^F-NOS kinetic measures

| ID* | M/F<sup>†</sup> | $V_T$<sup>‡</sup> | $K_1$<sup>§</sup> | $K_1/k_2$ | $BP_{ND}$<sup>¶</sup> | $vB$<sup>#</sup> |
| --- | --- | --- | --- | --- | --- | --- |
| <u>Electronic cigarette users (EC)</u> |  |  |  |  |  |  |
| EC-07 | F | 1.17 | 1.62 | 0.508 | 1.31 | 0.146 |
| EC-10 | M | 0.63 | 1.42 | 0.283 | 1.23 | 0.162 |
| EC-13 | F | 0.99 | 1.58 | 0.423 | 1.34 | 0.177 |
| EC-20 | M | 1.20 | 2.67 | 0.386 | 2.12 | 0.175 |
| EC-23 | M | 0.83 | 1.26 | 0.313 | 1.66 | 0.134 |
| <u>Combustible cigarette users (CIG)</u> |  |  |  |  |  |  |
| CIG-12 | M | 1.10 | 2.71 | 0.571 | 0.93 | 0.144 |
| CIG-14 | M | 4.74 | 2.29 | 3.417 | 0.39 | 0.142 |
| CIG-17 | M | 1.14 | 3.24 | 0.518 | 1.21 | 0.156 |
| CIG-22 | F | 1.45 | 2.70 | 0.663 | 1.29 | 0.110 |
| CIG-24 | F | 1.06 | 1.62 | 0.505 | 1.09 | 0.148 |
| <u>Never smoke/vape controls (CON)</u> |  |  |  |  |  |  |
| CON-01 | F | 1.15 | 1.95 | 0.579 | 0.98 | F |
| CON-03 | M | 1.04 | 1.85 | 0.480 | 1.17 | M |
| CON-05 | M | 0.91 | 1.37 | 0.401 | 1.27 | M |
| CON-06 | F | 1.53 | 3.19 | 0.742 | 1.07 | F |
| CON-09 | M | 1.18 | 1.62 | 0.555 | 1.13 | M |
| EC (n=5): |  | 0.966 ±0.240 | 1.710<br>±0.556 | 0.382 ±0.090 | 1.532 ±0.368 | 0.159 ±0.019 |
| CIG (n=5): |  | 1.897 ±1.595 | 2.512<br>±0.601 | 1.129 ±1.280 | 0.981 ±0.358 | 0.140 ±0.018 |
| CON (n=5): |  | 1.163 ±0.232 | 1.993<br>±0.702 | 0.552 ±0.127 | 1.123 ±0.109 | 0.156 ±0.021 |
| All (n=15): |  | 1.342 ±0.965 | 2.072<br>±0.671 | 0.688 ±0.765 | 1.212 ±0.371 | 0.152 ±0.020 |

^18^F-NOS BP_ND_ values differed significantly between groups, *H*(2) = 7.50, *p* = 0.02 (Figure 3). Post-hoc comparisons revealed that EC users had higher BP_ND_ values than cigarette smokers (*p* = 0.03) and healthy never smoke/vape controls (*p* = 0.01). Analysis of ^18^F-NOS V_T_ and K_1_ values showed no significant difference between groups (*p*’s > 0.09).

**Figure 3.**
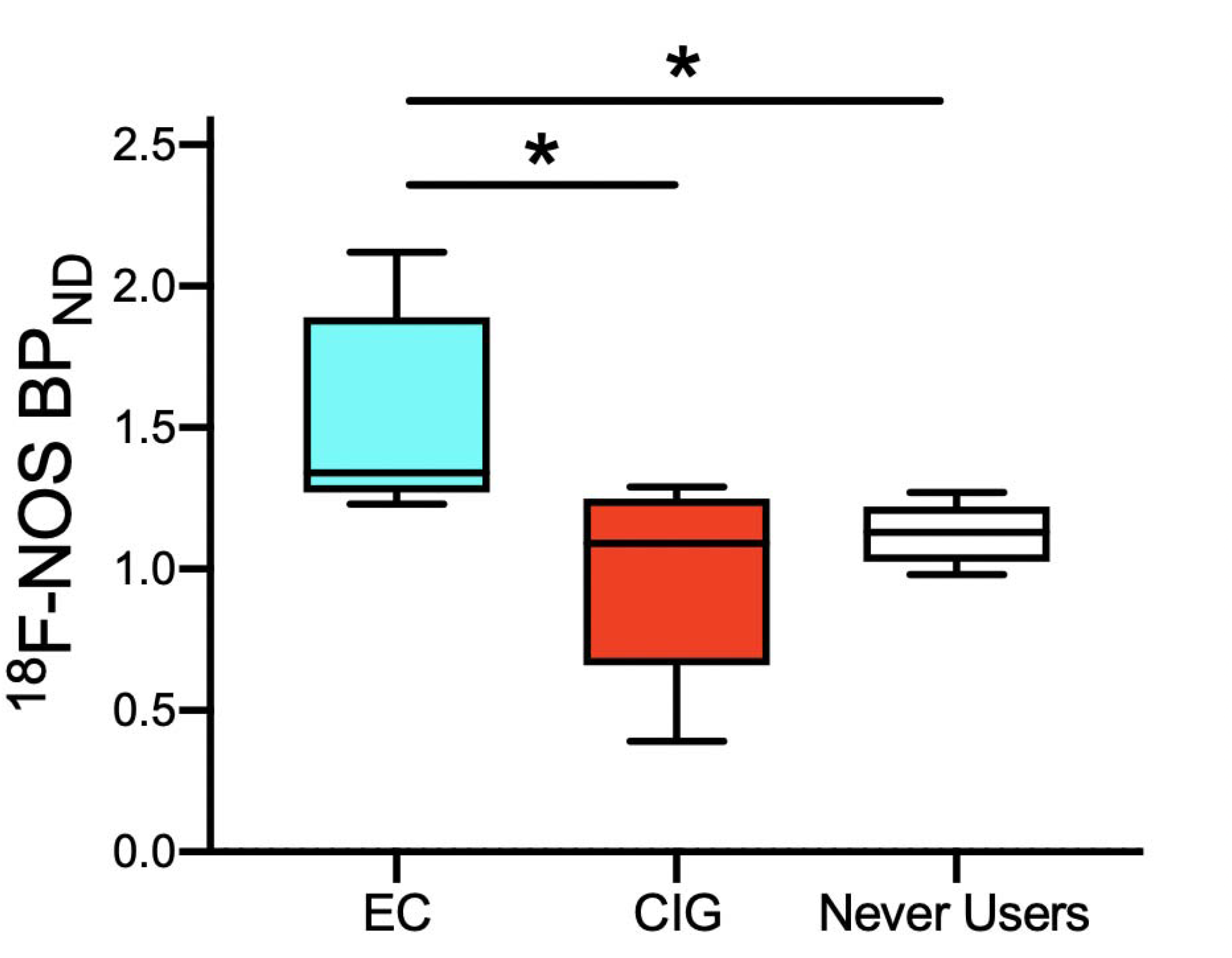
Boxplot of ^18^F-NOS BP_ND_ by Group. EC Users show higher ^18^F-NOS BPnd than never smoke/vape controls (*p* = 0.01) and cigarette smokers (*p* = 0.03). EC = E-Cigarette Users, CIG = Cigarette Smokers, Never Users = Never Smoke/Vape Controls. \**p* < 0.05

Groups did not significantly differ in plasma concentrations of proinflammatory cytokines (*p*’s > 0.16). We used Spearman’s rank-order correlations to examine the associations between daily smoking/vaping and proinflammatory cytokines and between imaging parameters and levels of proinflammatory cytokines. Among EC users only, there was a positive correlation between ^18^F-NOS BP_ND_ and TNF-α concentration (*r*_*s*_ = 0.87, *p* = 0.05). Among EC users and cigarette smokers, cigarettes per day and vaping episodes per day correlated with IL-6 levels (*r*_*s*_= 0.86, *p* = 0.006). No other correlations were statistically significant.

## DISCUSSION

EC use has increased dramatically, particularly among adolescents and young adults, and consequently, there is an urgent need for well-controlled studies of the effects of EC use on the lungs compared to those of cigarette smoking. To date, the existing literature is mainly comprised of *in vitro* and *ex vitro* cell culture studies or *in vivo* studies conducted in rodents, with a few studies of the effects of EC use on the human lung based on invasive approaches that do not assess the global burden of EC use on the lungs. This preliminary study addresses these gaps by using non-invasive, ^18^F-NOS PET lung imaging to quantify and compare lung inflammation in exclusive EC users, exclusive cigarette smokers, and never smoke/vape controls. Our preliminary ^18^F-NOS PET findings show that EC users have similar delivery of ^18^F-NOS to the lung tissue (K_1_) and iNOS availability (V_T_) similar to that of cigarette smokers and never smoke/vape controls. However, ^18^F-NOS BP_ND_ was significantly higher in the EC group than both cigarette smokers and controls. Moreover, ^18^F-NOS BP_ND_ in EC use was associated with the proinflammatory cytokine TNF-α. Cigarettes per day and vaping episodes per day correlated with IL-6 levels among cigarette smokers and EC users, respectively. To our knowledge, this is the first PET lung imaging study to demonstrate that EC users show a unique PET lung phenotype that is associated with known proinflammatory cytokines.

Although we did not see the expected increase in ^18^F-NOS uptake in cigarette smokers, our findings are consistent with recent work by Davis et al. (*38*) that used bronchoscopy to isolate alveolar macrophages from bronchoalveolar lavage samples in smokers, EC users, and never-smokers. These investigators found that EC use led to greater iNOS expression in alveolar macrophages than in smokers or nonsmokers. Animal and human studies show that iNOS expression is induced in most cell types upon exposure to inflammatory stimuli (*39*) and is associated with increased pulmonary NO (*40*). NO mediates neutrophil and macrophage actions that are thought to contribute to pulmonary oxidant stress and acute lung injury (*41*). Thus, our findings suggest that EC use may alter pulmonary oxidative stress responses and predispose to acute lung injury.

Although the groups showed similar levels of proinflammatory cytokines, EC users showed positive associations between ^18^F-NOS PET imaging parameters and TNF-α concentration. TNF-α is a proinflammatory cytokine produced by macrophages and secreted by neutrophil granulocytes at sites of injury (*42*) and is involved in the inflammatory cascade of acute lung injury (*43*). Indeed, studies show that proinflammatory cytokines induce iNOS expression in human alveolar cells in response to exposure to fine particulate matter (*44*). Consistent with our finding of greater ^18^F-NOS BP_ND_ in EC users, we suggest that these correlations provide additional evidence of the altered immune responses in the lungs of EC users.

Several limitations should be considered. First, we did not account for vaping topography, i.e., how an EC is used, including parameters such as puff duration, volume of puff, and duration of EC use episode, or EC device and power settings (e.g., battery voltage and coil resistance). These factors are important in differential exposure to nicotine and toxicants among e-cigarette users (*45*). While we used individually measured PET radiotracer parent fractions as a function of time to correct for the presence of radiolabelled metabolites in the blood, we could not separate lung ^18^F-NOS uptake due to binding of parent ^18^F-NOS from any binding of radiolabelled metabolites. Huang et al. asserted “because of [the metabolite’s] polarity, this metabolite is most likely excluded by the lung endothelium from entering the lung parenchyma” (*17*). Impacts of any lung binding of radiolabelled metabolites on estimates of ^18^F-NOS BP_ND_ will likely be inversely related to the validity of the Huang et al. assumption that polar ^18^F-NOS metabolites cannot penetrate lung endothelium. In addition, the sample sizes are small, so additional larger studies are needed to replicate these findings and provide greater statistical power for secondary analyses. Additional data collection is ongoing, as are studies assessing the effects of dual use (e.g., smoking and vaping) and the use of tetrahydrocannabinol (i.e., both combustible and e-liquid use).

### Conclusion

Using rigorous quantitative methods and a global technique to examine pulmonary oxidative stress, we find evidence that EC use causes a unique inflammatory response in the lungs, reflected both by PET measures of iNOS expression and proinflammatory cytokine concentrations. Future work is needed to elucidate fully the effect of EC use on respiratory health, especially the effects of chronic EC use.

## Data Availability

All data produced in the present study are available upon reasonable request to the authors.

## Acknowledgments

The study was supported by funding from the National Heart, Lung, and Brain Institute (R21 HL144673), the National Institute on Drug Abuse (P30 DA046345), the National Center for Advancing Translational Sciences of the National Institutes of Health under Award Number UL1TR001878, and in part by the Institute for Translational Medicine and Therapeutics’ (ITMAT) Transdisciplinary Program in Translational Medicine and Therapeutics.

## Key Points

QUESTION: What are the effects of e-cigarette use on pulmonary inflammation compared to combustible cigarette use and never smoking/vaping, as measured in vivo with ^18^F-NOS PET imaging?

PERTINENT FINDINGS: In this preliminary PET imaging study, e-cigarette users showed greater ^18^F-NOS non-displaceable binding potential (BP_ND_) than cigarette smokers and never smoke/vape controls. ^18^F-NOS BP_ND_ significantly correlated with the proinflammatory cytokine TNF-α in e-cigarette users. Additionally, when e-cigarette users and cigarette smokers were pooled together, vaping episodes/cigarettes per day correlated with IL-6 levels.

IMPLICATIONS FOR PATIENT CARE: Preliminary data indicate that e-cigarette users show a unique PET lung imaging phenotype associated with known proinflammatory cytokines, challenging the concept that e-cigarettes are a healthier alternative to cigarettes.

## References

1. WHO report on the global tobacco epidemic 2021: addressing new and emerging products. Geneva: World Health Organization; 2021. Licence: CC BY-NC-SA 3.0 IGO.

2. Wang TW, Gentzke AS, Neff LJ, et al. Disposable e-cigarette use among U.S. youth — An emerging public health challenge. N Engl J Med. 2021;384:1573–1576.

3. Miech R, Johnston L, O’Malley PM, Bachman JG, Patrick ME. Trends in adolescent vaping, 2017–2019. N Engl J Med. 2019;381:1490–1491.

4. Ioakeimidis N, Vlachopoulos C, Tousoulis D. Efficacy and safety of electronic cigarettes for smoking cessation: a critical approach. Hellenic J Cardiol. 2016;57:1–6.

5. Kalkhoran S, Glantz SA. E-cigarettes and smoking cessation in real-world and clinical settings: a systematic review and meta-analysis. Lancet Respir Med. 2016;4:116–128.

6. Pisinger C, Døssing M. A systematic review of health effects of electronic cigarettes. Prev Med. 2014;69:248–260.

7. Lundbäck B, Katsaounou P, Lötvall J. The up-rise in e-cigarette use – friend or foe? Respir Res. 2016;17:52, s12931-016-0371-2.

8. Cherian SV, Kumar A, Estrada-Y-Martin RM. E-Cigarette or vaping product-associated lung injury: a review. Am J Med. 2020;133:657–663.

9. Goniewicz ML, Smith DM, Edwards KC, et al. Comparison of nicotine and toxicant exposure in users of electronic cigarettes and combustible cigarettes. JAMA Netw Open. 2018;1:e185937.

10. Shields PG, Berman M, Brasky TM, et al. A review of pulmonary toxicity of electronic cigarettes in the context of smoking: a focus on inflammation. Cancer Epidemiol Biomark Prev. 2017;26:1175–1191.

11. Reidel B, Radicioni G, Clapp PW, et al. E-cigarette use causes a unique innate immune response in the lung, involving increased neutrophilic activation and altered mucin secretion. Am J Respir Crit Care Med. 2018;197:492–501.

12. Tong L, Sui Y, Jiang S, Yin Y. The association between lung fluorodeoxyglucose metabolism and smoking history in 347 healthy adults. J Asthma Allergy. 2021;14:301–308.

13. Sahota A, Naidu S, Jacobi A, et al. Atherosclerosis inflammation and burden in young adult smokers and vapers measured by PET/MR. Atherosclerosis. 2021;325:110–116.

14. Tateishi U, Hasegawa T, Seki K, Terauchi T, Moriyama N, Arai Y. Disease activity and 18F-FDG uptake in organising pneumonia: semi-quantitative evaluation using computed tomography and positron emission tomography. Eur J Nucl Med Mol Imaging. 2006;33:906–12.

15. Jones HA, Marino PS, Shakur BH, Morrell NW. In vivo assessment of lung inflammatory cell activity in patients with COPD and asthma. Eur Respir J. 2003;21:567–73.

16. Mostard RL, Verschakelen JA, van Kroonenburgh MJ, et al. Severity of pulmonary involvement and (18)F-FDG PET activity in sarcoidosis. Respir Med. 2013;107:439–47.

17. Huang HJ, Isakow W, Byers DE, et al. Imaging pulmonary inducible nitric oxide synthase expression with PET. J Nucl Med. 2015;56:76–81.

18. Hatori A, Yui J, Yamasaki T, et al. PET imaging of lung inflammation with [18F]FEDAC, a radioligand for translocator protein (18 kDa). PLoS One. 2012;7:e45065.

19. Branley HM, du Bois RM, Wells AU, Jones HA. Peripheral-type benzodiazepine receptors in bronchoalveolar lavage cells of patients with interstitial lung disease. Nucl Med Biol. 2007;34:553–8.

20. Notter T, Coughlin JM, Sawa A, Meyer U. Reconceptualization of translocator protein as a biomarker of neuroinflammation in psychiatry. Mol Psychiatry. 2018;23:36–47.

21. Koch A, Burgschweiger A, Herpel E, et al. Inducible NO synthase expression in endomyocardial biopsies after heart transplantation in relation to the postoperative course. Eur J Cardiothorac Surg. 2007;32:639–43.

22. Alderton WK, Cooper CE, Knowles RG. Nitric oxide synthases: structure, function and inhibition. Biochem J. 2001;357:593–615.

23. Smith KJ, Lassmann H. The role of nitric oxide in multiple sclerosis. Lancet Neurol. 2002;1:232–41.

24. Lechner M, Lirk P, Rieder J. Inducible nitric oxide synthase (iNOS) in tumor biology: the two sides of the same coin. Semin Cancer Biol. 2005;15:277–89.

25. Guo FH, De Raeve HR, Rice TW, Stuehr DJ, Thunnissen FB, Erzurum SC. Continuous nitric oxide synthesis by inducible nitric oxide synthase in normal human airway epithelium in vivo. Proc Natl Acad Sci U A. 1995;92:7809–13.

26. Khanna A, Guo M, Mehra M, Royal W. Inflammation and oxidative stress induced by cigarette smoke in Lewis rat brains. J Neuroimmunol. 2013;254:69–75.

27. Seimetz M, Parajuli N, Pichl A, et al. Inducible NOS inhibition reverses tobacco-smoke-induced emphysema and pulmonary hypertension in mice. Cell. 2011;147:293–305.

28. Chen GG, Lee TW, Xu H, et al. Increased inducible nitric oxide synthase in lung carcinoma of smokers. Cancer. 2008;112:372–381.

29. Bhandari V, Choo-Wing R, Chapoval SP, et al. Essential role of nitric oxide in VEGF-induced, asthma-like angiogenic, inflammatory, mucus, and physiologic responses in the lung. Proc Natl Acad Sci U A. 2006;103:11021–6.

30. Herrero P, Laforest R, Shoghi K, et al. Feasibility and dosimetry studies for 18F-NOS as a potential PET radiopharmaceutical for inducible nitric oxide synthase in humans. J Nucl Med. 2012;53:994–1001.

31. Zhou D, Lee H, Rothfuss JM, et al. Design and synthesis of 2-amino-4-methylpyridine analogues as inhibitors for inducible nitric oxide synthase and in vivo evaluation of [18F]6-(2-fluoropropyl)-4-methyl-pyridin-2-amine as a potential PET tracer for inducible nitric oxide synthase. J Med Chem. 2009;52:2443–53.

32. Kuntic M, Oelze M, Steven S, et al. Short-term e-cigarette vapour exposure causes vascular oxidative stress and dysfunction: evidence for a close connection to brain damage and a key role of the phagocytic NADPH oxidase (NOX-2). Eur Heart J. 2020;41:2472–2483.

33. Zigmond AS, Snaith RP. The Hospital Anxiety and Depression Scale. Acta Psychiatr Scand. 1983;67:361–370.

34. Foulds J, Veldheer S, Yingst J, et al. Development of a questionnaire for assessing dependence on electronic cigarettes among a large sample of ex-smoking E-cigarette users. Nicotine Tob Res. 2015;17:186–92.

35. Fagerstrom K. Determinants of tobacco use and renaming the FTND to the Fagerstrom Test for Cigarette Dependence. Nicotine Tob Res. 2012;14:75–8.

36. Kolthammer JA, Su K-H, Grover A, Narayanan M, Jordan DW, Muzic RF. Performance evaluation of the Ingenuity TF PET/CT scanner with a focus on high count-rate conditions. Phys Med Biol. 2014;59:3843–3859.

37. Holman BF, Cuplov V, Millner L, et al. Improved correction for the tissue fraction effect in lung PET/CT imaging. Phys Med Biol. 2015;60:7387–7402.

38. Davis ES, Ghosh A, Coakley RD, et al. Chronic E-Cigarette Exposure Alters Human Alveolar Macrophage Morphology and Gene Expression. Nicotine Tob Res. 2022;24:395–399.

39. Zamora R, Vodovotz Y, Billiar TR. Inducible Nitric Oxide Synthase and Inflammatory Diseases. Mol Med. 2000;6:347–373.

40. Mehta A, Guidot D. Alcohol and the lung. Alcohol Res Curr Rev. 2017;38:243–254.

41. Chow C-W, Herrera Abreu MT, Suzuki T, Downey GP. Oxidative stress and acute lung injury. Am J Respir Cell Mol Biol. 2003;29:427–431.

42. Tonstad S, Cowan JL. C-reactive protein as a predictor of disease in smokers and former smokers: a review. Int J Clin Pract. 2009;63:1634–1641.

43. Cross LJM, Matthay MA. Biomarkers in acute lung injury: insights into the pathogenesis of acute lung injury. Crit Care Clin. 2011;27:355–377.

44. Niu X, Ho KF, Hu T, et al. Characterization of chemical components and cytotoxicity effects of indoor and outdoor fine particulate matter (PM2.5) in Xi’an, China. Environ Sci Pollut Res. 2019;26:31913–31923.

45. Lee YO, Nonnemaker JM, Bradfield B, Hensel EC, Robinson RJ. Examining daily electronic cigarette puff topography among established and nonestablished cigarette smokers in their natural environment. Nicotine Tob Res. 2018;20:1283–1288.

